# What instructions are available to health researchers for writing lay summaries? A scoping review

**DOI:** 10.1101/2021.06.09.21258450

**Authors:** Karen M Gainey, Mary O’Keeffe, Adrian C Traeger, Danielle M Muscat, Christopher M Williams, Kirsten J McCaffrey, Steven J Kamper

**Affiliations:** Sydney Health Literacy Lab, School of Public Health, Faculty of Medicine and Health, The University of Sydney, Camperdown, Australia; Institute for Musculoskeletal Health, Sydney Local Health District and The University of Sydney, Camperdown, Australia; School of Medicine and Public Health, The University of Newcastle, Callaghan, Australia; University of Sydney, Sydney, Australia & Nepean Blue Mountains Local Health District, Nepean, Australia

**Author notes:** **Corresponding author:** Karen M Gainey - Edward Ford Building, School of Public Health, Faculty of Medicine and Health, The University of Sydney, Camperdown, NSW 2006, Australia.; Twitter: @KarenMGainey.

**Keywords:** Lay summary, plain-English summary, plain-language summary, health communication, health literacy

## Abstract

**Objective:** To better understand the characteristics of, and requirements for, lay summaries by reviewing journals, global health organisations, professional medical associations and multi-disciplinary organisations, consumer advocacy groups and funding bodies.

**Design:** Using a scoping review methodology, we searched the websites of each identified data source to determine if they require, suggest, or refer to lay summaries. Two reviewers extracted lay summary writing instructions from eligible data sources from Australia, USA, UK, Canada and New Zealand. Data sources were linked to the top 10 non-communicable diseases.

**Main Outcome Measures:** Using an inductive approach, we identified characteristics of lay summaries and lay summary writing instructions and extracted data on these characteristics. These characteristics are lay summary formats, audience, requirements, authorship and labels, and elements of lay summary writing instructions (e.g. word count/length). We also noted who was expected to write the lay summaries, whether they were mandatory or optional, and the terms used for to denote them.

**Results:** The websites of 526 data sources were searched. Of these, 124 published or mentioned lay summaries and 108 provided writing instructions. For lay summaries, most were in journals, written by the author of the published paper, and only half were mandatory. Thirty-three distinct labels for a lay summary were identified, the most common being “graphical abstract”, “highlights” and “key points”. From the lay summary writing instructions, the most common elements for written lay summaries referred to: structure (86%), content (80%) and word count/length (74%). The least common elements were readability (3%), use of jargon, acronyms and abbreviations (24%), and wording (29%). The target audience was unclear in 68 of 108 (63.0%) of lay summary instructions.

**Discussion:** Although we identified over 100 sources provided instructions for writing lay summaries, very few provided instructions related to readability, use of jargon, acronyms and abbreviations, and wording. Some instructions provided structured formats via subheadings or questions to guide content, but not all. Only half mandated the use of lay summaries.

**Conclusion:** For lay summaries to be effective, writing instructions should consider the intended audience, ideally incorporating consumer input into their development. Presently, lay summaries are likely to be inaccessible to many consumers, written at a high reading level, with jargon, acronyms and abbreviations. Ideally, all research articles will have an accompanying lay summary. Mandatory lay summaries, however, are of limited value without clear and thorough instructions to guide authors.

**Public and patient involvement statement:** Patients or the public were not involved in the design, or conduct, or reporting, or dissemination plans of our research study.

**Protocol and registration:** We conducted a scoping review using methods outlined in the PRISMA extension for scoping reviews and information in the Joanna Briggs Institute Reviewers’ Manual for scoping reviews. A protocol for this study was completed prior to data analysis and is on Open Science Framework.

## INTRODUCTION

Most health research is not written with the public in mind as it contains jargon and acronyms and is usually written at a high reading level.(1) Lay summaries are condensed summaries of research articles written in plain, easy-to-understand language aimed at a non-scientific audience.(1) Ideally, lay summaries contain no jargon or technical language.(2) This makes them an ideal tool for disseminating reliable health information to the public.(3)

Scientists are accustomed to describing their work using jargon and technical concepts and may find it difficult to describe their research in a way that is accessible to a non-expert lay audience.(2) To guide authors, many journals and organisations provide instructions for writing lay summaries. The level of guidance they provide varies, however, and the advice is not always clear and thorough.(4) Despite the availability of instructions for writing lay summaries, research suggests that most lay summaries are written at a high reading level, which may make them difficult for consumers to understand.(5,6) There is no accepted consensus document to guide the writing of lay summaries, so authors must determine for themselves how best to proceed in writing the ideal lay summary for their paper.

Lay summaries are an important means of disseminating research findings to the public.(1) Unless non-experts can easily understand lay summaries, they may misinterpret the messages they contain.(1) Instructions should guide the author to write the lay summary with the intended audience in mind by avoiding jargon and ensuring the readability level is appropriate, for example by recommending a readability tool.

Research on instructions for writing lay summaries is scarce. Two previous reviews have assessed the content of lay summaries and writing instructions provided by a range of journals in the fields of biology, economics, medicine(7) and biomedicine.(8) One review focused primarily on lay summary writing instructions from consumer advocacy groups and included one list of instructions from a scientific paper and one published by a university.(9) The most comprehensive study of lay summary writing instructions was a review conducted and published by the journal *eLife*. This review included over fifty data sources such as journals and scientific organisations.(10) However, the only element of lay summary writing instructions noted by this *eLife* review was word count/length.(10)

Reporting guidelines have been developed to support researchers in implementing complex processes, providing transparency, accuracy and consistency to the reporting of health research.(11,12) Research guidelines have been used since 1996 when the Consolidated Standard of Reporting Trials (CONSORT) statement was developed, and examples includes the Preferred Reporting Items for Systematic Reviews and Meta-analysis (PRISMA) and Grading of Recommendations Assessment, Development and Evaluation (GRADE) guidelines.(11) When these guidelines are applied, the methodological and reporting quality of studies improves.(11) Use of these guidelines has increased over time as authors and publishers become more familiar with them.(11) A detailed analysis of lay summary writing instructions currently provided to authors will establish a foundation upon which we can gain a better understanding of the development of lay summaries and ways to enhance their value as a communication tool. As lay summaries become a more common part of health research communication, the development of consistent guidelines for writing lay summaries will be necessary.

The aim of this scoping review is to better understand the characteristics of lay summaries by reviewing journals, global health organisations, professional medical associations and multi-disciplinary organisations, consumer advocacy groups and funding bodies. We compiled the lay summary writing instructions from these sources and expect that the results from this review will assist in the development of consistent and uniform guidelines for writing lay summaries.

## METHODS

### Protocol and registration

We conducted a scoping review using methods outlined in the PRISMA extension for scoping reviews(13) and information in the Joanna Briggs Institute Reviewers’ Manual for scoping reviews.(14) A protocol for this study was completed prior to data analysis and is on Open Science Framework.(15)

### Data sources

We searched the following data sources: journals, global health organisations, professional medical organisations and multi-disciplinary associations, consumer advocacy groups and health research funding bodies.

### Eligibility criteria

Eligibility criteria were developed for each data source (Table 1). Within these data sources, we only included those which included research based on the top ten non-communicable diseases (NCDs) responsible for the greatest global disability burden,(16) most of which are chronic health conditions. We chose this focus because people with chronic health conditions are frequent users of these health information sources.(17,18) For professional medical organisations and multi-disciplinary associations, consumer advocacy groups and health research funding bodies, we included those from Australia, USA, UK, Canada and New Zealand. We chose these five countries as they are largely English speaking and have ready access to these sources. Examples include The Cardiac Society of Australia and Diabetes UK.

**Table 1.** Data sources and eligibility criteria.

| **Data source** | **Definition of data source** | **Eligibility criteria** | **Search date(s)**<br>**(YYYY-MM-DD)** |
| --- | --- | --- | --- |
| Journals | Peer-reviewed health and medical journals | Based on Web of Science categories from Journal of Citation Reports:<br>- Top 50 Medicine, General and Internal<br>- Top 25 from each of: Primary Health Care, Cardiac and Cardiovascular System, Rheumatology, Behavioural Sciences, Respiratory System and Clinical Neurology, | 2019-03-05 |
| Global health organisations | International health bodies or agencies relevant to the top ten NCDs that: 1.synthesise and disseminate health and medical information for researchers, clinicians and the public, or 2. specialise in clinical evidence/knowledge summaries | Initial list based on author consensus and consultation with colleagues: The World Health Organisation (WHO), Centers for Disease Control and Prevention, Cochrane Collaboration, Campbell Collaboration, Joanne Briggs Institute & National Institute for Health and Care Excellence.<br>All WHO partners whose projects linked to one of the top 10 NCDs and were active after the year 2000.<br>Advanced Google keyword searches using keywords such as 'global health organisation'. First ten pages were reviewed (see Appendix A). | Google searches on 2019-08-06<br>WHO Partnership & Collaborative list updated May 2019 |
| Professional medical associations or multidisciplinary organisations | Professional associations for groups of health and medical practitioners | Associations affiliated with the journals identified in the journal search.<br>Organisations associated with professions specific to the top ten NCDs in Australia, USA, UK, Canada and New Zealand.<br>Advanced Google keyword searches. First two pages per NCD per country were reviewed (see Appendix A). | 2019-06-10 & 2019-06-11 |
| Consumer advocacy groups | Groups that provide information and support to consumers with health conditions | Groups associated with health conditions represented by the top ten NCDs in Australia, USA, UK, Canada and New Zealand.<br>Advanced Google keyword searches. First two pages per NCD per country were reviewed (see Appendix A). | 2019-06-11 to 2019-06-16 |
| Funding bodies | National government bodies that fund health and medical research | Peak health research funding bodies for Australia, USA, UK, Canada and New Zealand. Advanced Google keyword search. First two pages of Google search per country were reviewed (see Appendix A). | 2019-06-17 |

### Search

The full search strategy and date(s) for each data source is detailed in Table 1. One reviewer conducted all Google searches based on the search strings in Appendix A. The relevant web pages were saved to PDF and the cache and browsing data cleared between each search.

### Selection of data sources

Two reviewers independently viewed all Google search results and selected relevant websites. When selected websites were agreed upon, both reviewers visited each website and extracted all data items. Discussion resolved all disagreements.

### Data charting process

A data charting form was developed by one researcher containing the key study characteristics to extract. Two reviewers searched the website of each identified data source to determine if they require, suggest, or refer to lay summaries. They then extracted lay summary writing instructions from the data sources. These instructions formed the primary data for this study. We also collected information on several categories of observations and recorded the data items according to these category components.

For journals, both reviewers viewed two original articles in each journal and noted whether a lay summary was included. If so, the article was saved as a PDF. We then consulted the author instructions and searched for the lay summary writing instructions. We saved these instructions for data analysis.

For all other data sources, both reviewers searched the website of each data source for any mention of lay summaries. Where available, reviewers used the search function, searching common terms for a lay summary such as “lay summary”, “plain English summary”, “plain language summary” and “summary”. We also searched for lay summaries under each tab on the website home page. When lay summaries and instructions for writing them were located, we followed the same process we followed when searching journals.

The most concise and widely used definition of a lay summary is that developed by the INVOLVE group, which was established in 1996 as part of the National Institute for Health to facilitate public involvement in research.(19) According to jargon buster produced by INVOLVE, a lay summary is “a brief summary of a research project or a research proposal that has been written for members of the public, rather than researchers or professionals. It should be written in plain English, avoid the use of jargon and explain any technical terms that have to be included”.(19) We took a broader definition of ‘lay summary’ for this review, including any summary or distillation of a research study that was not the abstract. This broader definition helped ensure we captured all available lay summaries. During data charting, we located lay summaries and instructions that suggested the target audience for the summary was not a non-expert lay public. We identified three clear target audiences for lay summaries. (Table 2).

**Table 2.**
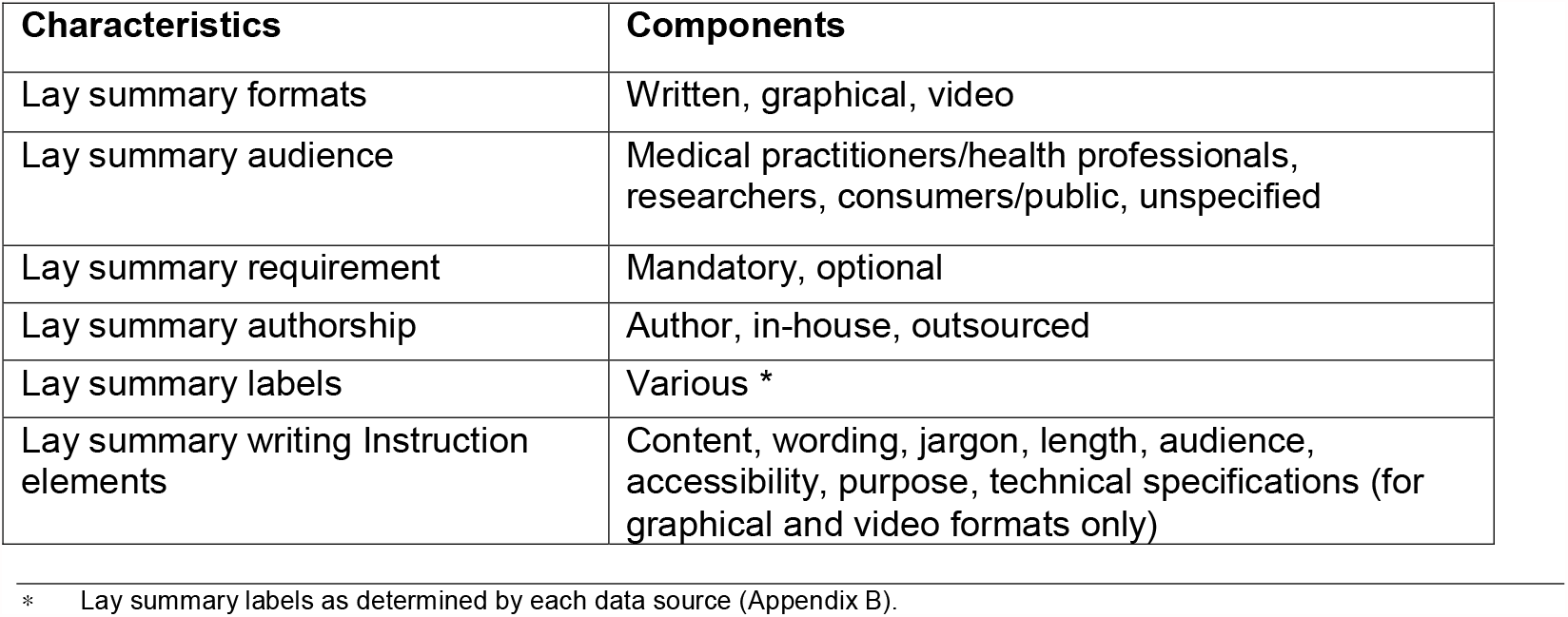
Characteristics and components of lay summaries and lay summary writing instructions.

| Characteristics | Components |
| --- | --- |
| Lay summary formats | Written, graphical, video |
| Lay summary audience | Medical practitioners/health professionals, researchers, consumers/public, unspecified |
| Lay summary requirement | Mandatory, optional |
| Lay summary authorship | Author, in-house, outsourced |
| Lay summary labels | Various * |
| Lay summary writing Instruction elements | Content, wording, jargon, length, audience, accessibility, purpose, technical specifications (for graphical and video formats only) |

\* Lay summary labels as determined by each data source (Appendix B).

### Data items

Using an inductive approach, we identified characteristics of lay summaries and lay summary writing instructions and extracted data on these characteristics. (Table 2).We located three formats for lay summaries: written, graphical and video summaries. Written lay summaries are the most well-known format, however journal publishers now produce graphical and video summaries, referred to by the labels graphical abstract and video abstract. In this review, we refer to all three formats as lay summaries for consistency.

### Synthesis of results

We collated results for the lay summaries and the lay summary writing instructions in the review. We summarised the number of websites that mentioned or required a lay summary and those that provided lay summary writing instructions, grouped by data source. Data was also extracted for; format of the lay summary, the audience for the lay summary, requirement for a lay summary (optional or mandatory), the party expected to write the lay summary, and all labels used to refer to the lay summaries.

## RESULTS

### Main findings

Figure 1 shows the process of website selection. Overall, 556 websites were identified across the five data sources, of which 526 were searched for the study. We excluded 30 websites because of duplication, broken website links or inability to access necessary text. Remaining websites included 179 journals, 41 global health organisations, 147 professional medical associations and multidisciplinary organisations, 154 consumer advocacy groups and five funding bodies. From these 526 websites, 124 (23.6%) published or mentioned lay summaries and 108 (20.5%) provided lay summary writing instructions. (Table 3). Of note, none of the data sources we identified detailed how their instructions were developed or mentioned end-user involvement.

**Figure 1.**
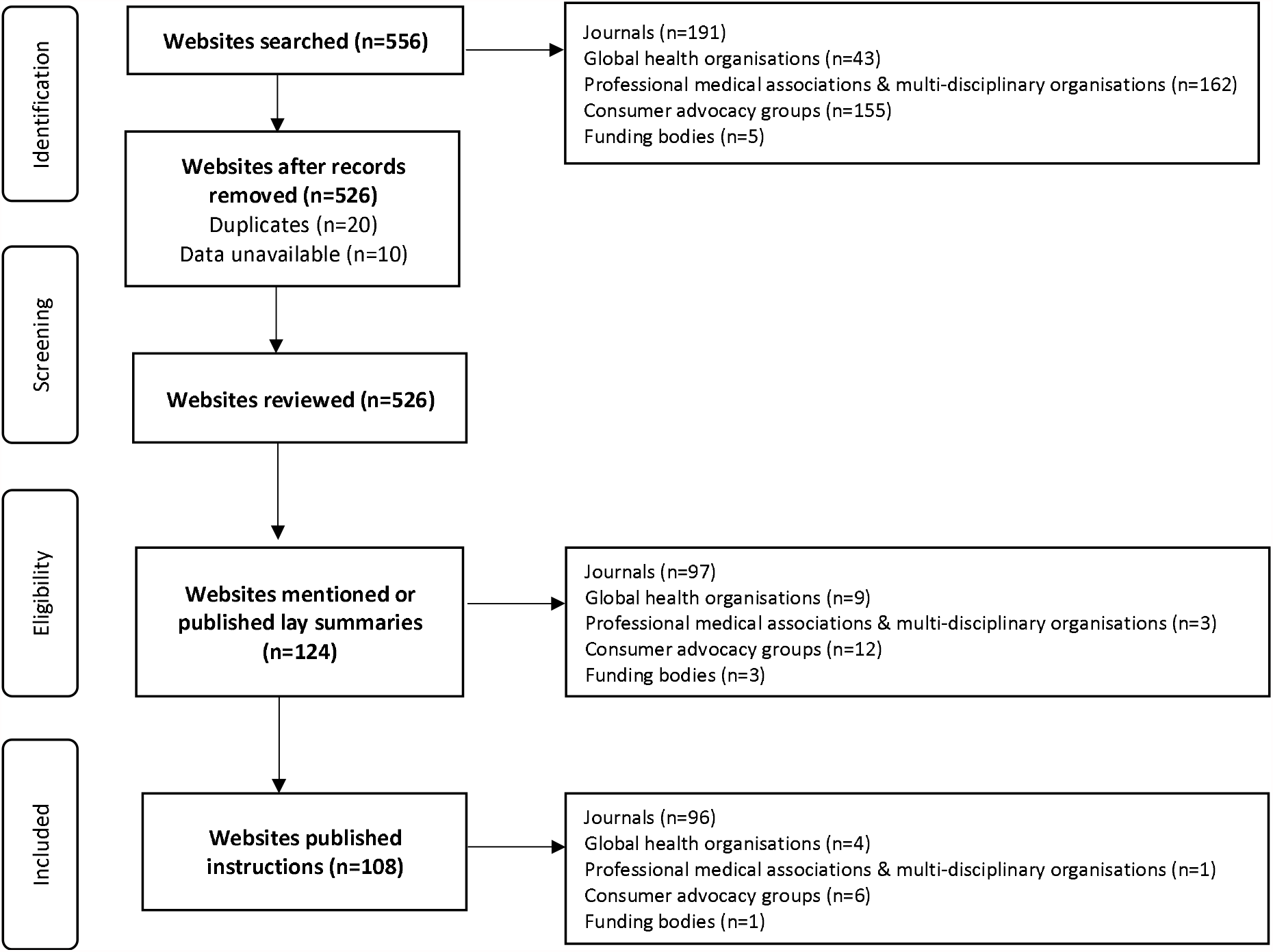
Flow diagram of included data sources

**Table 3.** Overview of findings.

| **Data source** | **Number** | **Produced/mentioned lay summaries** | **Provided lay summary writing instructions** |
| --- | --- | --- | --- |
| Journals | 179 | 98 (54.7%) | 97 (54.2%) |
| Global health organisations | 41 | 8 (19.5%) | 3 (7.3%) |
| Professional medical associations & multi-disciplinary organisations | 147 | 3 (2.0%) | 1 (0.68%) |
| Consumer advocacy groups | 154 | 12 (7.8%) | 6 (3.9%) |
| Funding bodies | 5 | 3 (60%) | 1 (20%) |
| **Totals** | **526** | **124 (23.6%)** | **108 (20.5%)** |

### Lay summaries

Journals contributed the largest number of lay summaries and lay summary writing instructions, with only half being mandatory. Most lay summaries were written by the author of the published paper.

#### Format of lay summaries

Lay summaries were produced in three formats: written, graphical and video. Of the 108 writing instructions for lay summaries, 95 (88.0%) were for lay summaries in a written format, 36 (33.3%) for a graphical format and 13 (12.0%) for a video format. The instructions for most graphical and video lay summaries appeared besides those for written lay summaries.

#### Audience for lay summaries

From all data sources, we identified three target audiences for lay summaries: medical practitioners/health professionals, researchers, consumers/public, the first two of which are considered an expert audience. The target audience was not specified, not clear or covered several categories in 68 of 108 (63.0%) of lay summary instructions. Forty (37.0%) writing instructions specified a target audience for the summary; 28/108 (25.9%) for an expert audience and 12/108 (11.1%) for consumers/public. For those summaries targeted at an expert audience, 10/108 (9.3%) were for medical practitioners/health professionals and 18/108 (16.7%) for researchers.

#### Requirement for lay summaries

Of the 124 sources that mentioned or published lay summaries, 63 (50.8%) considered lay summaries mandatory at least some article types e.g. original research, reviews and basic science papers. In 61 (49.2%) instances, lay summaries were optional or not specified. Lay summaries were mandatory for seven consumer advocacy groups and an important part of funding applications. These include the Prostate Cancer Foundation of Australia and the British Heart Foundation.

#### Authorship of lay summaries

On 104 websites (96.3%) the authors of the article were required to write the lay summary. Four (3.7%) summaries were written by an in-house team; one journal, two global health organisations and one professional medical association and multidisciplinary organisation. We did not locate any data sources that out-sourced the writing of their lay summaries. Wiley (journal publishers of seven included journals), however, offers a professional lay summary service to authors for a fee.

#### Labels for lay summaries

Thirty-three distinct labels for a lay summary were identified, accounting for 164 lay summaries. Some journals published multiple lay summaries, for example, a written and either graphical or video summary. The most common labels were ‘Graphical Abstracts’ (*Journal of Heart and Lung Transplantation*–journal), ‘Highlights’ (*American Journal of Medicine* - journal) and ‘Key Points’ (*JAMA Cardiology*–journal). These labels were used 37 (22.6%), 27 (16.5%) and 22 (13.4%) times respectively. Of the remaining thirty labels for a lay summary, 19 (11.6%) were found only once in our review. In two instances, no label was identified (Appendix B).

The label ‘lay summary’ was often used by global health organisations and consumer advocacy groups, but only three journals. The label ‘plain-language summary’ was only used by the Cochrane and Campbell Collaborations. None of the data sources used the label ‘plain-English summary’.

### Lay summary writing Instructions

For each format of lay summary, we identified key elements in the instructions such as content, purpose and structure (Table 4). The most common elements for written lay summaries referred to: structure (84.2%), content (84.2%) and word count/length (77.9%). The least common elements were readability (3.2%), use of jargon, acronyms and abbreviations (25.3%), and wording (30.5%). All instructions for graphical lay summaries referred to content and technical specifications, with (69.2%) including purpose. Most instructions for video abstracts included technical specifications (92.3%).

**Table 4.**
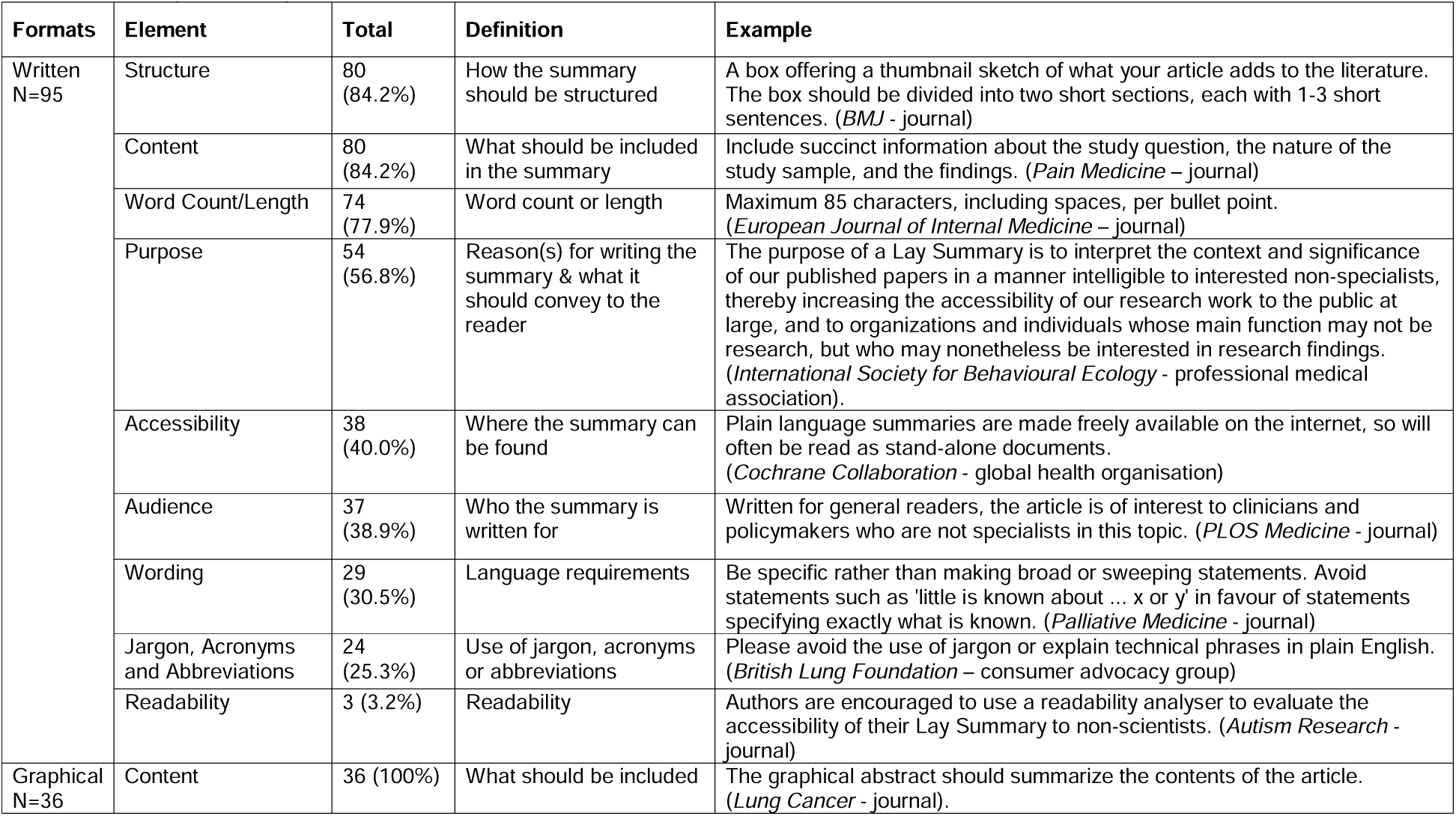
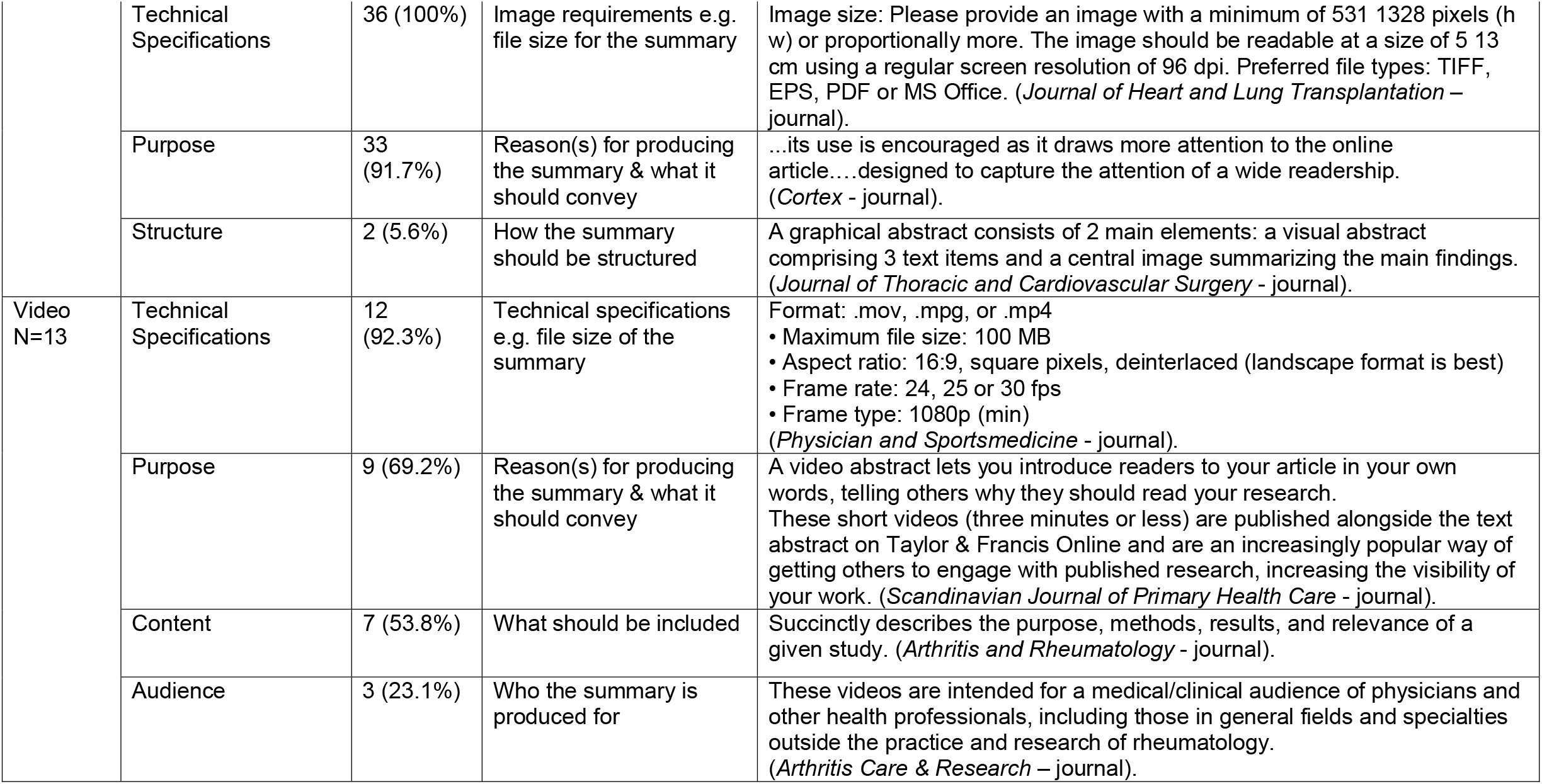
Lay summary instructions.

| Formats | Element | Total | Definition | Example |
| --- | --- | --- | --- | --- |
| Written<br>N=95 | Structure | 80<br>(84.2%) | How the summary should be structured | A box offering a thumbnail sketch of what your article adds to the literature. The box should be divided into two short sections, each with 1-3 short sentences. (*BMJ* - journal) |
|  | Content | 80<br>(84.2%) | What should be included in the summary | Include succinct information about the study question, the nature of the study sample, and the findings. (*Pain Medicine* – journal) |
|  | Word Count/Length | 74<br>(77.9%) | Word count or length | Maximum 85 characters, including spaces, per bullet point. (*European Journal of Internal Medicine* – journal) |
|  | Purpose | 54<br>(56.8%) | Reason(s) for writing the summary & what it should convey to the reader | The purpose of a Lay Summary is to interpret the context and significance of our published papers in a manner intelligible to interested non-specialists, thereby increasing the accessibility of our research work to the public at large, and to organizations and individuals whose main function may not be research, but who may nonetheless be interested in research findings. (*International Society for Behavioural Ecology* - professional medical association). |
|  | Accessibility | 38<br>(40.0%) | Where the summary can be found | Plain language summaries are made freely available on the internet, so will often be read as stand-alone documents. (*Cochrane Collaboration* - global health organisation) |
|  | Audience | 37<br>(38.9%) | Who the summary is written for | Written for general readers, the article is of interest to clinicians and policymakers who are not specialists in this topic. (*PLOS Medicine* - journal) |
|  | Wording | 29<br>(30.5%) | Language requirements | Be specific rather than making broad or sweeping statements. Avoid statements such as 'little is known about ... x or y' in favour of statements specifying exactly what is known. (*Palliative Medicine* - journal) |
|  | Jargon, Acronyms and Abbreviations | 24<br>(25.3%) | Use of jargon, acronyms or abbreviations | Please avoid the use of jargon or explain technical phrases in plain English. (*British Lung Foundation* – consumer advocacy group) |
|  | Readability | 3 (3.2%) | Readability | Authors are encouraged to use a readability analyser to evaluate the accessibility of their Lay Summary to non-scientists. (*Autism Research* - journal) |
| Graphical<br>N=36 | Content | 36 (100%) | What should be included | The graphical abstract should summarize the contents of the article. (*Lung Cancer* - journal). |
|  | Technical Specifications | 36 (100%) | Image requirements e.g. file size for the summary | Image size: Please provide an image with a minimum of 531 1328 pixels (h w) or proportionally more. The image should be readable at a size of 5 13 cm using a regular screen resolution of 96 dpi. Preferred file types: TIFF, EPS, PDF or MS Office. (*Journal of Heart and Lung Transplantation – journal*). |
|  | Purpose | 33 (91.7%) | Reason(s) for producing the summary & what it should convey | ...its use is encouraged as it draws more attention to the online article....designed to capture the attention of a wide readership. (*Cortex - journal*). |
|  | Structure | 2 (5.6%) | How the summary should be structured | A graphical abstract consists of 2 main elements: a visual abstract comprising 3 text items and a central image summarizing the main findings. (*Journal of Thoracic and Cardiovascular Surgery - journal*). |
| Video<br>N=13 | Technical Specifications | 12 (92.3%) | Technical specifications e.g. file size of the summary | Format: .mov, .mpg, or .mp4<br>• Maximum file size: 100 MB<br>• Aspect ratio: 16:9, square pixels, deinterlaced (landscape format is best)<br>• Frame rate: 24, 25 or 30 fps<br>• Frame type: 1080p (min)<br>(*Physician and Sportsmedicine - journal*). |
|  | Purpose | 9 (69.2%) | Reason(s) for producing the summary & what it should convey | A video abstract lets you introduce readers to your article in your own words, telling others why they should read your research. These short videos (three minutes or less) are published alongside the text abstract on Taylor & Francis Online and are an increasingly popular way of getting others to engage with published research, increasing the visibility of your work. (*Scandinavian Journal of Primary Health Care - journal*). |
|  | Content | 7 (53.8%) | What should be included | Succinctly describes the purpose, methods, results, and relevance of a given study. (*Arthritis and Rheumatology - journal*). |
|  | Audience | 3 (23.1%) | Who the summary is produced for | These videos are intended for a medical/clinical audience of physicians and other health professionals, including those in general fields and specialties outside the practice and research of rheumatology. (*Arthritis Care & Research – journal*). |

The instructions for writing lay summaries varied in content, intended audience and level of detail, ranging from a single sentence to several paragraphs. For 15 (12.1%) lay summaries, no instructions were provided at all. Large commercial publishing companies such as Elsevier often had standard instructions that applied to each of their journals.

Instructions for how to structure the lay summary most commonly referred to the use of bullet points and/or the use of specific headings such as ‘The Known, The New, The Implications’ (*Medical Journal of Australia* - journal). Instructions for content varied in length and level of detail, often as questions to address or headings to use, for example, ‘What is already known about this subject?, What does this study add?, How might this impact on clinical practice?’, (*Heart* - journal). Instructions for word count/length were usually expressed as a maximum number of words or sentences per bullet point. Word counts ranged from 10 to 500 words. Bullet points varied from 3 to 6 bullet points of 85 characters or 15 words each. Technical specifications for both graphical and video abstracts were detailed and precise, specifying file size, format and resolution.

## DISCUSSION

### Summary of evidence

In this scoping review of journals, global health organisations, professional medical organisations and multi-disciplinary associations, consumer advocacy groups and funding bodies from Australia, USA, UK, Canada and New Zealand, we found that only 23.6% published or mentioned lay summaries, with just over 100 providing instructions for writing lay summaries of health research. Very few provided instructions related to readability, use of jargon, acronyms and abbreviations, and wording. Some instructions provided structured formats via subheadings or questions to guide content, but not all. Only half mandated the use of lay summaries.

### Strengths and limitations

This review represents the most comprehensive review of lay summary writing instructions, including data sources across 526 websites. This is the first review to synthesise information on the style, content and technical specifications of instructions for producing lay summaries. However, our results were limited to information that was freely available and accessible on each website. Health and medical publishing is dominated by a few large publishing companies. Therefore, many of the instructions for writing lay summaries we identified were common across multiple journals, which may influence the proportion of elements in the instructions. However, we selected journals across seven categories from the journal of citation ranking reports. The journals included in this review represent high quality, widely read journals.

### Comparison to the literature

This work builds on the existing evidence-base for lay summaries to date. A review of the biomedical literature showed the labels commonly used to describe a lay summary are ‘lay summary’, ‘plain-language summary’ and ‘plain-English summary’.(1,3-8,20-22) The INVOLVE definition of lay summaries(2) also uses the label ‘plain-English summary’. These labels, however, are contrary to the findings of this review, which showed much greater variation in labels used for lay summaries and identified only 18 (11.0%) data sources that used these common labels. This is concerning given that the use of varied and imprecise labels for lay summaries may cause these summaries to not reach their target audience; the identification of a lay summary by the reader often begins with its label, and an unfamiliar or confusing label may deter readers. A universally accepted, clear and well-defined label may increase appeal of the lay summary and engagement by readers. Use of labels more easily accepted or recognised would also enable search engines to more easily locate lay summaries for users, making lay summaries more useful.(21)

In addition to the labels given to lay summaries, this review also builds on the literature related to the content of instructions for writing such summaries. Only 3% of instructions identified in this review recommended the use of a readability tool, and fewer than one-third of instructions gave direction regarding type of language and/or jargon. This finding is inconsistent with the INVOLVE definition of lay summaries which states that lay summaries “should be written clearly and simply, without jargon and with an explanation of any technical terms”.(2) Specifying readability is likely to be important because previous research has showed that most lay summaries are written at a high reading level, which may make them difficult for consumers to understand.(5,6) Poor readability can hinder the ability of a non-expert lay audience to improve their knowledge and understanding of their health condition.(23) This can lead to misinformation and poor self-management, potentially resulting in reduced health status.(23) The solution is to provide health information, such as lay summaries that are “accurate, accessible, and readable”.(18)

### Implications for research and practice

Our review highlights the need for instructions to specify the intended audience of the lay summary. Some labels reflect their intended audience (e.g. “clinical perspective”, “clinical significance” for health practitioners/clinicians). Other labels, however, were ambiguous and may confuse readers, particularly the lay public (e.g. “Context”, “What’s New”, “Highlights” and “Summary at a Glance”). If the target audience and label is unclear, it may not be obvious that it is referring to a lay summary of the research article. Future work should explore the labelling preferences of each target audience, and work should be done to facilitate greater consistency across data sources.

Related to this, we took a definition of lay summaries broader in scope than that of INVOLVE(2) to ensure this was a comprehensive review. Despite this broader definition, we did not expect to locate as many as a quarter of lay summaries specifically targeted at an expert audience (medical practitioners/health professionals and researchers). The *eLife* review(10) noted two journals also include research summaries aimed at an expert audience e.g. The *Proceedings of the National Academy of Sciences*. In 2016, the journal *Nature* trialled the use of summaries written specifically for other researchers.(10) With the presence of article abstracts, it is unclear why some publishers of health research focus on delivering research summaries to an expert audience at the expense of conveying health research to a non-expert lay public. Given the technical nature of most research articles, there is a need for lay summaries aimed at a lay audience to satisfy their demand for trustworthy health information.(20) The public, especially those with chronic illnesses represented by the top 10 NCDs(16), consider journals a primary source of health information.(20) Lay summaries are also a tool that a non-expert lay public can use with their treating clinicians to aid in shared-decision making.(20)

### Unanswered questions and future research

This review has highlighted two primary issues with lay summary writing instructions. First, inconsistency of guidance for authors writing lay summaries. Second, many don’t appear to appreciate the needs of a non-expert lay audience; our findings suggest that distributors of research evidence may prioritise the structure, content and length of the lay summary over accessibility for the target audience. One solution to these issues would be the development of a framework for writing lay summaries that is universally accepted by publishers of research evidence. This framework could include the easy identification as a lay summary by its label, consideration for the target audience (e.g. grade reading level and identifying the target audience at the top of the lay summary) and describe requirements of consumer collaboration. Given that most lay summaries (96%) are written by the authors of the research article who are unlikely to have had formal training on writing for a non-expert lay audience,(23) providing authors with training along with a standard framework could help them write more effective lay summaries. The skills and techniques used to write a research article are different to those required to communicate in lay language.(23)

None of the data sources detailed how their instructions were developed or mentioned end-user involvement. Involving consumers in the co-design process could produce lay summaries that better meet their needs.(24) Few guidelines exist to help authors work with stakeholders such as consumers on the development of lay summaries, however the protocol developed for researchers in the AGE-WELL network is a promising foundation.(1) This protocol outlines six steps in co-creating lay summaries with stakeholders. These steps begin with an investigation of the principles for writing a lay summary, and the target audience and key stakeholders. This is followed by involving stakeholders in writing lay summaries and conducting workshops to provide them with information and support.(1) Guidelines and lay summary instructions, applied correctly, can lead to high-quality research for dissemination to a wide audience. To assist their ongoing implementation, collaboration between authors, publishers and other stakeholders would be beneficial.(1)

A final area for future research relates to graphic abstracts. Graphical abstracts can attract attention to an article and are an ideal format for sharing on social media, which could improve their reach to a wider audience.(25) However, they need to be well designed with an emphasis on the main findings, which takes time and effort.(25) In 89% of graphical and 75% of video lay summaries respectively, we found the purpose was clearly outlined as attracting reader attention and increasing engagement. An evaluation of these two formats for lay summaries would be beneficial to gauge their effectiveness in meeting this objective. If effective, these formats for lay summaries offer an important alternative to written lay summaries for readers for whom a more visual medium is easier to use.

## CONCLUSION

We discovered that most instructions for lay summaries referred to structure, content and word count/length. Elements such as avoiding jargon, acronyms and abbreviations and readability were notably absent from most instructions. For lay summaries to be effective, writing instructions should specify the intended audience. As a primary end-user, consumer input into the development of lay summaries could improve accessibility of lay summaries for their intended audience. Without consumer input, lay summaries are likely to be inaccessible to many consumers, written at a high reading level, with jargon, acronyms and abbreviations. The requirement for all research articles to have an accompanying lay summary would aid in exposing a non-expert lay public to health research. However, mandatory lay summaries are of limited value unless authors are provided with clear and thorough instructions that help authors write lay summaries with the target audience in mind.

## Supporting information

PRISMA ScR Checklist

## Data Availability

Additional detailed data can be obtained from the authors on request.
The corresponding author confirms that the manuscript is an honest, accurate, and transparent account of the study being reported. No important aspects of the study have been omitted and any discrepancies from the study as planned have been disclosed.

## Contributorship statement

KG, SK, AT and CW conceptualised the research question and developed the study protocol. KG and MO’K conducted data charting with any disagreements resolved by SK. KG conducted the data analysis under the supervision of SK, DM and KMcC. KG drafted the manuscript with all authors contributing to revisions.

## Funding

not applicable.

## Ethical approval

Not required.

## Competing interests

All authors have completed the ICMJE uniform disclosure form at http://www.icmje.org/coi_disclosure.pdf. Two authors declare financial support from the National Health and Medical Research Council (NHMRC) of Australia for contribution to their fellowships and one author declares financial support from an NHMRC program grant (APP1113532) entitled ‘Using healthcare wisely: reducing inappropriate use of tests and treatments’. NHMRC funding was not related to this study however this organisation might have an interest in the submitted work. All authors declare no other financial relationships with any other organisation that might have an interest in the submitted work in the previous three years; no other relationships or activities that could appear to have influenced the submitted work.

## Data sharing

Additional detailed data can be obtained from the authors on request. The corresponding author confirms that the manuscript is an honest, accurate, and transparent account of the study being reported. No important aspects of the study have been omitted and any discrepancies from the study as planned have been disclosed.

## Dissemination to participants and related patient and public communities

No patients were involved in setting the research question or the outcome measures, nor were they involved in the design and implementation of the study. We plan to disseminate the study findings to the general public through social media, our bi-monthly newsletter and through partnerships with relevant organisations.

## Acknowledgements

This review was presented as a virtual poster at the What Works Global Summit, 2020, Integrated Health Literacy Annual Research Conference and Health Literacy in Action conference, 2020 Institute for Health Advancement 19^h^ Annual Health Literacy Conference 2020 and the CHF Virtual Summit, March 2021. It was also presented as a lightening talk at the Evidence and Implementation Summit, 2021

