## Appendices for "What instructions are available to health researchers for writing lay summaries? A scoping review"

**Appendix A**

Unless otherwise specified, all search strings are typed into the box “all these words” in the advanced Google search option. For all searches, we will set the region to “any region” and the language to “English”.

Global Health Organisations

Search strings used in Google for Global Health Organisations are as follows:

1. global health organisations [simple search option]
2. international health organisation [simple search option]
3. global health organisation OR evidence OR knowledge OR research OR synthesis
4. global health organisation OR evidence OR knowledge OR research OR summary
5. international health organisation OR evidence OR knowledge OR research OR synthesis
6. international health organisation OR evidence OR knowledge OR research OR summary

Professional Medical Organisations and Multidisciplinary Associations

Search strings for Professional Medical Organisations and Multidisciplinary Associations are as follows:

1. cardiac Australia medical association OR organisation OR institute
2. cancer Australia medical OR association OR organisation OR institute
3. musculoskeletal Australia medical OR association OR organisation OR institute
4. mental Australia medical OR association OR organisation OR institute
5. respiratory Australia medical OR association OR organisation OR institute
6. neurological Australia medical OR association OR organisation OR institute
7. diabetes Australia medical OR association OR organisation OR institute
8. kidney Australia” medical OR association OR organisation OR institute
9. digestive Australia medical OR association OR organisation OR institute
10. blind Australia medical OR association OR organisation OR institute
11. deaf Australia medical OR association OR organisation OR institute
12. cardiac United States of America USA medical OR association OR organisation OR institute
13. cancer United States of America USA medical OR association OR organisation OR institute
14. musculoskeletal United States of America USA medical OR association OR organisation OR institute
15. mental United States of America USA medical OR association OR organisation OR institute
16. respiratory United States of America USA medical OR association OR organisation OR institute
17. neurological United States of America USA medical OR association OR organisation OR institute
18. diabetes United States of America USA medical OR association OR organisation OR institute
19. kidney United States of America USA medical OR association OR organisation OR institute
20. digestive United States of America USA medical OR association OR organisation OR institute
21. deaf United States of America USA medical OR association OR organisation OR institute
22. blind United States of America USA medical OR association OR organisation OR institute
23. cardiac United Kingdom UK medical OR association OR organisation OR institute
24. cancer United Kingdom UK medical OR association OR organisation OR institute
25. musculoskeletal United Kingdom UK medical OR association OR organisation OR institute
26. mental United Kingdom UK medical OR association OR organisation OR institute
27. respiratory United Kingdom UK medical OR association OR organisation OR institute
28. neurological United Kingdom UK medical OR association OR organisation OR institute
29. diabetes United Kingdom UK medical OR association OR organisation OR institute
30. kidney United Kingdom UK medical OR association OR organisation OR institute
31. digestive United Kingdom UK medical OR association OR organisation OR institute
32. deaf United Kingdom UK medical OR association OR organisation OR institute
33. blind United Kingdom UK medical OR association OR organisation OR institute
34. cardiac Canada medical OR association OR organisation OR institute
35. cancer Canada medical OR association OR organisation OR institute
36. musculoskeletal Canada medical OR association OR organisation OR institute
37. mental Canada medical OR association OR organisation OR institute
38. respiratory Canada medical OR association OR organisation OR institute
39. neurological Canada medical OR association OR organisation OR institute
40. diabetes Canada medical OR association OR organisation OR institute
41. kidney Canada medical OR association OR organisation OR institute
42. digestive Canada medical OR association OR organisation OR institute
43. deaf Canada medical OR association OR organisation OR institute
44. blind Canada medical OR association OR organisation OR institute
45. cardiac New Zealand NZ medical OR association OR organisation OR institute
46. cancer New Zealand NZ medical OR association OR organisation OR institute
47. musculoskeletal New Zealand NZ medical OR association OR organisation OR institute
48. mental New Zealand NZ medical OR association OR organisation OR institute
49. respiratory New Zealand NZ medical OR association OR organisation OR institute
50. neurological New Zealand NZ medical OR association OR organisation OR institute
51. diabetes New Zealand NZ medical OR association OR organisation OR institute
52. kidney New Zealand NZ medical OR association OR organisation OR institute
53. digestive New Zealand NZ medical OR association OR organisation OR institute
54. deaf New Zealand NZ medical OR association OR organisation OR institute
55. blind New Zealand NZ medical OR association OR organisation OR institute

Consumer Advocacy Groups

Search strings for Consumer Advocacy Groups are as follows:

1. consumer OR patient OR advocacy OR support OR group OR organisation cardiac Australia
2. consumer OR patient OR advocacy OR support OR group OR organisation cancer Australia
3. consumer OR patient OR advocacy OR support OR group OR organisation musculoskeletal Australia
4. consumer OR patient OR advocacy OR support OR group OR organisation mental health Australia
5. consumer OR patient OR advocacy OR support OR group OR organisation respiratory Australia
6. consumer OR patient OR advocacy OR support OR group OR organisation neurological Australia
7. consumer OR patient OR advocacy OR support OR group OR organisation diabetes Australia
8. consumer OR patient OR advocacy OR support OR group OR organisation kidney Australia
9. consumer OR patient OR advocacy OR support OR group OR organisation digestive Australia
10. consumer OR patient OR advocacy OR support OR group OR organisation deaf Australia
11. consumer OR patient OR advocacy OR support OR group OR organisation blind Australia
12. consumer OR patient OR advocacy OR support OR group OR organisation cardiac United States of America USA
13. consumer OR patient OR advocacy OR support OR group OR organisation cancer United States of America USA
14. consumer OR patient OR advocacy OR support OR group OR organisation musculoskeletal United States of America USA
15. consumer OR patient OR advocacy OR support OR group OR organisation mental United States of America USA
16. consumer OR patient OR advocacy OR support OR group OR organisation respiratory United States of America USA
17. consumer OR patient OR advocacy OR support OR group OR organisation neurological United States of America USA
18. consumer OR patient OR advocacy OR support OR group OR organisation diabetes United States of America USA
19. consumer OR patient OR advocacy OR support OR group OR organisation kidney United States of America USA
20. consumer OR patient OR advocacy OR support OR group OR organisation digestive United States of America USA
21. consumer OR patient OR advocacy OR support OR group OR organisation deaf United States of America USA
22. consumer OR patient OR advocacy OR support OR group OR organisation blind United States of America USA
23. consumer OR patient OR advocacy OR support OR group OR organisation cardiac United Kingdom UK
24. consumer OR patient OR advocacy OR support OR group OR organisation cancer United Kingdom UK
25. consumer OR patient OR advocacy OR support OR group OR organisation musculoskeletal United Kingdom UK
26. consumer OR patient OR advocacy OR support OR group OR organisation mental United Kingdom UK
27. consumer OR patient OR advocacy OR support OR group OR organisation respiratory United Kingdom UK
28. consumer OR patient OR advocacy OR support OR group OR organisation neurological United Kingdom UK
29. consumer OR patient OR advocacy OR support OR group OR organisation diabetes United Kingdom UK
30. consumer OR patient OR advocacy OR support OR group OR organisation kidney United Kingdom UK
31. consumer OR patient OR advocacy OR support OR group OR organisation digestive United Kingdom UK
32. consumer OR patient OR advocacy OR support OR group OR organisation deaf United Kingdom UK
33. consumer OR patient OR advocacy OR support OR group OR organisation blind United Kingdom UK
34. consumer OR patient OR advocacy OR support OR group OR organisation cardiac Canada
35. consumer OR patient OR advocacy OR support OR group OR organisation cancer Canada
36. consumer OR patient OR advocacy OR support OR group OR organisation musculoskeletal Canada
37. consumer OR patient OR advocacy OR support OR group OR organisation mental Canada
38. consumer OR patient OR advocacy OR support OR group OR organisation respiratory Canada
39. consumer OR patient OR advocacy OR support OR group OR organisation neurological Canada
40. consumer OR patient OR advocacy OR support OR group OR organisation diabetes Canada
41. consumer OR patient OR advocacy OR support OR group OR organisation kidney Canada
42. consumer OR patient OR advocacy OR support OR group OR organisation digestive Canada
43. consumer OR patient OR advocacy OR support OR group OR organisation deaf Canada
44. consumer OR patient OR advocacy OR support OR group OR organisation blind Canada
45. consumer OR patient OR advocacy OR support OR group OR organisation cardiac New Zealand NZ
46. consumer OR patient OR advocacy OR support OR group OR organisation cancer New Zealand NZ
47. consumer OR patient OR advocacy OR support OR group OR organisation musculoskeletal New Zealand NZ
48. consumer OR patient OR advocacy OR support OR group OR organisation mental New Zealand NZ
49. consumer OR patient OR advocacy OR support OR group OR organisation respiratory New Zealand NZ
50. consumer OR patient OR advocacy OR support OR group OR organisation neurological New Zealand NZ
51. consumer OR patient OR advocacy OR support OR group OR organisation diabetes New Zealand NZ
52. consumer OR patient OR advocacy OR support OR group OR organisation kidney New Zealand NZ
53. consumer OR patient OR advocacy OR support OR group OR organisation digestive New Zealand NZ
54. consumer OR patient OR advocacy OR support OR group OR organisation deaf New Zealand NZ
55. consumer OR patient OR advocacy OR support OR group OR organisation blind New Zealand NZ

Funding Bodies

Search strings for Funding Bodies are as follows:

1. national government medical research funding organisation OR association Australia
2. national government medical research funding organisation OR association United States of America USA
3. national government medical research funding organisation OR association United Kingdom UK
4. national government medical research funding organisation OR association Canada
5. national government medical research funding organisation OR association New Zealand NZ

**Appendix B**

**Labels for lay summaries**

| **Label for lay summary** | **Frequency** | **Data source** | | | | |
| --- | --- | --- | --- | --- | --- | --- |
|  |  | **Journals** | **Global health organisations** | **Professional medical associations & multidisciplinary organisations** | **Consumer advocacy groups** | **Funding bodies** |
| Graphical Abstract | 37 | 36 | 1 | 0 | 0 | 0 |
| Highlights | 27 | 27 | 0 | 0 | 0 | 0 |
| Key Points | 22 | 20 | 0 | 1 | 0 | 1 |
| Lay Summary | 15 | 3 | 2 | 1 | 9 | 0 |
| Video Abstract | 12 | 12 | 0 | 0 | 0 | 0 |
| Key Message | 10 | 7 | 1 | 1 | 0 | 1 |
| Translational Perspective | 4 | 4 | 0 | 0 | 0 | 0 |
| Clinical Perspective | 3 | 3 | 0 | 0 | 0 | 0 |
| Plain Language Summary | 3 | 1 | 2 | 0 | 0 | 0 |
| Research in Context | 3 | 3 | 0 | 0 | 0 | 0 |
| What is already known on this topic & What this topic adds | 3 | 3 | 0 | 0 | 0 | 0 |
| Unclear | 2 | 2 | 0 | 0 | 0 | 0 |
| Summary Box | 2 | 0 | 2 | 0 | 0 | 0 |
| What’s New | 2 | 2 | 0 | 0 | 0 | 0 |
| At a Glance Commentary | 1 | 1 | 0 | 0 | 0 | 0 |
| Article Summary | 1 | 1 | 0 | 0 | 0 | 0 |
| Author Summary | 1 | 1 | 0 | 0 | 0 | 0 |
| Brief Commentary | 1 | 1 | 0 | 0 | 0 | 0 |
| Central Message | 1 | 1 | 0 | 0 | 0 | 0 |
| Clinical Significance | 1 | 1 | 0 | 0 | 0 | 0 |
| Context | 1 | 0 | 1 | 0 | 0 | 0 |
| How This Fits In | 1 | 1 | 0 | 0 | 0 | 0 |
| Key Findings | 1 | 0 | 0 | 0 | 1 | 0 |
| Key Questions | 1 | 1 | 0 | 0 | 0 | 0 |
| Key Statements | 1 | 1 | 0 | 0 | 0 | 0 |
| Lay Abstract | 1 | 0 | 0 | 0 | 0 | 1 |
| Perspective Statement | 1 | 1 | 0 | 0 | 0 | 0 |
| Research Lay Summary | 1 | 0 | 0 | 0 | 1 | 0 |
| Research Summary | 1 | 0 | 0 | 0 | 1 | 0 |
| Statement of Significance | 1 | 1 | 0 | 0 | 0 | 0 |
| Summaries for Patients | 1 | 1 | 0 | 0 | 0 | 0 |
| Summary at a Glance | 1 | 1 | 0 | 0 | 0 | 0 |
| The Known, The New, The Implications | 1 | 1 | 0 | 0 | 0 | 0 |
